# Incorporating data from multiple endpoints in the analysis of clinical trials: example from RSV vaccines

**DOI:** 10.1101/2023.02.07.23285596

**Authors:** Ottavia Prunas, Joukje E. Willemsen, Louis Bont, Virginia E. Pitzer, Joshua L. Warren, Daniel M. Weinberger

## Abstract

**Background:** To achieve licensure, interventions typically must demonstrate efficacy against a primary outcome in a randomized clinical trial. However, selecting a single primary outcome *a priori* is challenging. Incorporating data from multiple and related outcomes might help to increase statistical power in clinical trials. Inspired by real-world clinical trials of interventions against respiratory syncytial virus (RSV), we examined methods for analyzing data on multiple endpoints.

**Method:** We simulated data from three different populations in which the efficacy of the intervention and the correlation among outcomes varied. We developed a novel permutation-based approach that represents a weighted average of individual outcome test statistics (*varP*) to evaluate intervention efficacy in a multiple endpoint analysis. We compared the power and type I error rate of this approach to two alternative methods: the Bonferroni correction (*bonfT*) and another permutation-based approach that uses the minimum P-value across all test statistics (*minP*).

**Results:** When the vaccine efficacy against different outcomes was similar, *VarP* yielded higher power than *bonfT* and *minP;* in some scenarios the improvement in power was substantial. In settings where vaccine efficacy was notably larger against one endpoint compared to the others, all three methods had similar power.

**Conclusions:** Analyzing multiple endpoints using a weighted permutation method can increase power while controlling the type I error rate in settings where outcomes share similar characteristics, like RSV outcomes. We developed an R package, *PERMEATE*, to guide selection of the most appropriate method for analyzing multiple endpoints in clinical trials.

## Introduction

To receive licensure from a regulatory body like the FDA, investigators need to demonstrate that new vaccines are efficacious in a Phase 3 randomized controlled trial. Phase 3 trials have become increasingly complex and costly in recent years [1]. By definition, clinical trials expose participants to experimental vaccines that have potential benefits but also have potential safety issues. For both of these reasons, trials need to be designed to have the greatest probability of success in the most efficient way.

Phase 3 trials typically must specify one or more primary endpoints, and the ultimate success or failure of the trial hinges on the ability to demonstrate a significant reduction in the occurrence of the primary endpoint among those receiving the intervention. The primary endpoint needs to be carefully selected based on clinical relevance, ability of the vaccine to prevent the outcome, and the incidence rate in the study population. When there are multiple possible clinically relevant outcomes that are related, it is particularly difficult to determine which endpoint should be designated as “primary”. Incorporating data from multiple endpoints (or ‘outcomes’) in evaluations of vaccine efficacy could provide a more complete understanding of the effects of the intervention and increase the statistical power for detecting a true effect [2]. In general, there are two main advantages to including multiple endpoints in the primary analysis: (i) to increase the power of statistical tests and thereby reduce the required sample size of the trial, and (ii) to describe treatment effects more comprehensively, specifically for diseases where a single outcome measure does not fully represent the treatment effect [3]. Therefore, analyzing multiple endpoints instead of a single endpoint could potentially avert wasted research effort.

As an example of the pitfalls of selecting a single clinical endpoint, a maternal RSV vaccine failed to achieve licensure in 2020 because it did not show efficacy against the primary endpoint of medically significant RSV lower respiratory tract infection (LRTI) [4]. The trial only slightly missed the threshold for statistical significance [4]. However, the vaccine did show efficacy against related secondary endpoints, including hospitalization with RSV LRTI. If the analysis plan of the maternal RSV vaccine had incorporated data from multiple endpoints, it may have been reasonable to conclude that the vaccine was efficacious [4]. However, the rigidity of the regulatory process led to this potentially useful vaccine being largely abandoned.

Choosing the best approach to analyze data on multiple endpoints in clinical trials depends on the objectives and design of the trial, as well as the characteristics of the endpoints themselves [5]. When analyzing multiple endpoints, it is important to continue to control the type I error rate (i.e., the probability of falsely rejecting the null hypothesis of no vaccine effect when it is true). The most common approach for doing so is to analyze each endpoint independently and adjust the corresponding P-values using the Bonferroni correction [6, 7]. However, this approach does not account for the overlap or correlation between endpoints, which typically leads to overly conservative P-values and lower power compared to other approaches [2, 8, 9]. For instance, most hospitalizations with RSV LRTI are considered medically significant, but not all medically significant RSV LRTI will be hospitalized. Resampling methods, such as permutation approaches, could provide more power when there is overlap between endpoints, while at the same time maintaining control of the overall type I error rate [2, 3, 5, 7, 10]. However, despite the increasing use of multiple endpoints in randomized clinical trials, guidance is still limited on how to analyze and interpret the resulting data [2, 11].

Here, we propose and evaluate a novel approach for analyzing data on multiple endpoints where we derive a permutation-based test statistic that represents a weighted average of individual outcome test statistics. We compare the power and type I error rate of this approach with two established methods using simulated data that have different vaccine effect sizes and levels of correlation among the endpoints, mirroring real-world RSV clinical trials. Additionally, we analyze multiple endpoint data from a real-world clinical trial using these methods and compare the findings. Our aim is to provide practical recommendations on the preferred method based on the specific setting, along with tools for implementing the approaches we describe.

## Methods

We compare the performance of three methods for analyzing multiple endpoints across different simulated scenarios. The first method, the Bonferroni correction (*bonfT*), is one of the most common approaches for analyzing multiple outcomes due to its simplicity in application. The second and third methods are permutation-based approaches, which allow us to flexibly combine the test statistics from multiple outcomes into a single metric for making statistical inference. For all methods, we used the risk ratio (RR) as our test statistic for the individual endpoints, which is directly connected to vaccine efficacy (i.e., vaccine efficacy = 1 - RR).

### Bonferroni correction

A common approach for analyzing data on multiple endpoints is to first analyze each outcome independently. In this setting, we use a one-sided two-sample test of proportions comparing the attack rate in the intervention and control arms to obtain P-values for each endpoint. To correct for the multiple hypothesis tests being performed, *bonfT* divides the original statistical significance level (i.e., *α* level, here set to 0.05) by the total number of outcomes (i.e., *M*). The method considers a treatment effect to be statistically significant if any of the endpoint-specific P-values are less than *α*/*M*.

### Permutation-based methods

Permutation methods estimate the distribution of a selected test statistic when the null hypothesis is true by repeatedly “shuffling” the treatment group labels for individuals in the control and intervention groups in the observed data. The test statistic is then calculated for each permuted dataset by comparing the prevalence of the outcome in the “control” and “intervention” groups when these labels are randomly assigned. The actual test statistic from the observed data (i.e., with correct group labels) is compared to this null distribution; a P-value is then calculated based on the quantile where the actual test statistic falls on the null distribution (Figure S1).

#### Test statistics

For each permuted dataset, the minimum P-value approach (*minP*) first computes a P-value for each endpoint based on the individually analyzed RRs and then selects the smallest P-value as its test statistic [12]. Similar to *bonfT*, we used a one-sided two-sample test of proportions to obtain the P-values for each endpoint. The test statistic for a single permuted dataset is given as 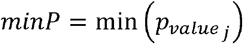 for outcome *j* ∈ {1, …, *M*}.

Alternatively, we developed a novel approach where we pooled the RRs from each outcome together using a weighted average (*varP*). Specifically, we calculated the RR for each outcome and created a summary test statistic by multiplying each RR (on the log scale) by a factor proportional to the inverse of its variance, such that the test statistic is given as:

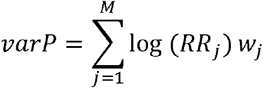

where 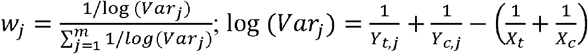 and *Y*_*c,j*_ are the number of cases of endpoint *j* in the treatment and control arms, respectively; and *X*_*t*_ and *X*_*c*_ are the number of individuals in the treatment and control arms, respectively.

We tested additional approaches by varying how the test statistic was constructed; in one case, we used the product of RRs as our test statistic; in another case, we used a summary test statistic where each RR was multiplied by the weighted average of the outcomes in the control group. However, these methods did not perform well.

#### Creating the null distribution

We randomly shuffled the treatment and control labels for the individuals *N*=999 times to obtain a distribution of the permuted test statistics for both *minP* and *varP*. In parallel, we computed the same test statistics for the observed data, deriving *minP* _*obs*_ and *varP* _*obs*_.

The permutation P-value is defined as the proportion of permuted datasets that have a test statistic equal to or more extreme than that of the observed data. This corresponds to a one-tailed test (Figure S1 in the supplementary file). Specifically, the P-values are defined as 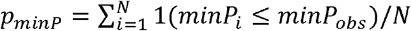 and 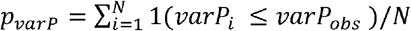 for the two test statistics, respectively. The statistical significance level was set at 0.05 for these tests (i.e., *α* = 0.05).

### Data simulation

To test the performance of the three different methods for analyzing multiple endpoints, we performed a study in which 10,000 different datasets were simulated and analyzed with each method. We considered several scenarios in which we varied the number of individuals in the control and treatment groups, the expected incidence of each outcome in the treatment group, and the effect of the intervention against each outcome (as quantified by the risk ratio). For each scenario, we varied the correlation among outcomes between 0.01 and 0.8.

Individual-level binary outcomes for each endpoint (i.e. whether or not the outcome occurred during the follow-up period of the trial) were independently generated using the R package *bindata* [13], which first simulates a correlated vector of continuous responses based on treatment status (e.g., vaccination status) and then dichotomizes those responses as above/below zero to create the binary outcome. For example, when considering two endpoints, the vector of binary response for individual 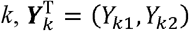, was simulated as *Y*_*kj*_ = 1(*β*_0*j*_ + *β*_1*j*_ *z*_*k*_ + *ϵ*_*kj*_ > 0), *j* = 1,2, where z_*k*_ indicates whether participant *k* received the intervention (z_*k*_=1) or control (z_*k*_=0), *β*_0*j*_ specifies the proportion of the control group that experiences the outcome of interest during the follow-up period, *β*_1*j*_ is the intervention effect for outcome *j*, and *ϵ*_*kj*_ are correlated errors simulated from a multivariate Gaussian distribution, such as 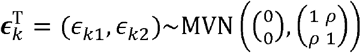, with *ρ* representing the correlation coefficient. Correlation in the latent continuous variables results in correlation between the binary responses specific to an individual, regardless of treatment status. We selected *β*_0*j*_ and *β*_1*j*_ parameters so that the incidence in the control and treatment arms was *λ*_*c,j*_ and *λ*_*t,j*_, respectively (i.e., the proportion of participants in the control and treatment arms experiencing the outcome of interest). This model was extended to simulate three outcomes [2].

We assumed each person could develop the *M*=3 different outcomes, with the degree of correlation among the outcomes varied between 0.01 and 0.8, *ρ* ∈ {0.01, 0.2, 0.4, 0.6, 0.8}. Also, our approach assumed all-or-nothing vaccine protection. After simulating the data, we computed the observed RR for each outcome as 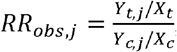 with *X*_*t*_, *X*_*c*_, *X*_*t,j*_ and *Y*_*c*.*j*_ previously defined.

We simulated data based on three different scenarios, inspired by three published clinical trials of RSV prophylaxis and vaccines. The MAKI trial was a study of the effect of palivizumab, a monoclonal antibody used for RSV prophylaxis, for prevention of childhood wheeze (scenario A)[14]; the Melody trial was a phase III trial testing the effectiveness of extended half-life RSV prophylaxis, nirsevimab, against lower respiratory disease in infants (scenario B) [15]; and the Prepare trial was a phase III trial of a maternal vaccine, RSV-F, for the prevention of RSV-associated lower respiratory disease in infants (scenario C) [4].

In the “MAKI-type” simulated data, we assumed a similar number of participants in each arm, with *X*_*t*_ = *X*_*c*_ = 200, high incidence of the three outcomes in the control arm, i.e. the proportion of participants in the control arm experiencing the outcome of interest (*λ*_*cj*_ ∼ 0.12-0.22), similar effect sizes of the intervention against the three outcomes (*RR*_*j*_ ∼ 60-70%), and expected number of cases equal to 20-50 for the three outcomes [14]. The “Melody-type” data had a 2:1 ratio between individuals randomized to the treatment and control arms, with *X*_*t*_ = 996 and *X*_*c*_ = 496, low incidence of the three outcomes (*λ*_*cj*_ ∼ 0.02-0.05), corresponding to 10-50 expected cases, and heterogeneous effect sizes (*RR*_*j*_ ∼ 25-60%) [15]. Finally, the “Prepare-type” data had more participants than the other two trials, with *X*_*t*_ = 2,765 and *X*_*c*_ = 1,430, low incidence of the three outcomes (*λ*_*cj*_ ∼ 0.01-0.04), corresponding to 10-40 expected cases, and comparable effect sizes against the various outcomes (*RR*_*j*_ ∼ 50-60%); in the latter setting, the intervention had the strongest effect size against the outcome with the lowest incidence [4]. Details of the different simulated scenarios are summarized in Table 1.

**Table 1:** Simulated scenarios. Three scenarios were tested with different sample size, effect size, and incidence. For all scenarios, the correlation among the outcomes was varied between *ρ*=0.01 and *ρ*=0.80.

| Scenario | Sample size<br>number of<br>participants in the<br>control ($X_c$) and<br>treatment ($X_t$)<br>groups | Incidence of each<br>outcome in the<br>control arm<br>($\lambda_{c,j}$) | Intervention<br>effect size<br>risk ratio against<br>outcome $j$ ($RR_j$) | Example<br>outcomes for<br>RSV trials |
| --- | --- | --- | --- | --- |
| Scenario A) High<br>and comparable<br>effect size and<br>incidence (MAKI-type data) | $X_c = X_t = 200$ | $\lambda_{c,1}=0.22,$<br>$\lambda_{c,2}=0.20,$<br>$\lambda_{c,3}=0.12$ | $RR_1=0.60,$<br>$RR_2=0.60,$<br>$RR_3=0.70$ | Childhood asthma,<br>wheeze, and use of<br>asthma<br>medications |
| Scenario B) Low<br>incidence and<br>heterogeneous<br>effect size<br>(Melody-type data) | $X_c = 496, X_t = 994$ | $\lambda_{c,1}=0.05,$<br>$\lambda_{c,2}=0.02,$<br>$\lambda_{c,3}=0.03$ | $RR_1=0.25,$<br>$RR_2=0.40,$<br>$RR_3=0.60$ | Medically attended<br>RSV LRTI,<br>hospitalization for<br>RSV LRTI, and<br>hospitalization for<br>any RSV in<br>children |
| Scenario C) Low<br>incidence and high<br>effect size<br>(Prepare-type data) | $X_c = 1,430,$<br>$X_t = 2,765$ | $\lambda_{c,1}=0.02,$<br>$\lambda_{c,2}=0.04,$<br>$\lambda_{c,3}=0.01$ | $RR_1=0.60,$<br>$RR_2=0.55,$<br>$RR_3=0.50$ | Medically<br>significant RSV<br>LRTI, RSV LRTI<br>with severe<br>hypoxemia, and<br>hospitalization for<br>RSV LRTI |

### Power and type I error

We computed the power of each method as the proportion of the 10,000 simulations for which a significant effect was detected, as previously defined for each method. Similarly, we computed the type I error by setting *RR*_*j*_=1 for each outcome and then calculating the proportion of datasets for which we incorrectly found a significant effect.

### The MAKI trial case study: RSV prevention and asthma in healthy preterm infants

We also tested the different analytical approaches against real-world clinical trial data from the MAKI trial. MAKI was a double-blind, randomized, placebo-controlled trial involving 429 infants born at 32-35 weeks of gestation recruited during their first year of life between 2008-2010 [14, 16]. Participants were randomized to receive palivizumab prophylaxis or placebo. The trial reported a causal relationship between prevention of RSV infection during infancy and reduced frequency of subsequent wheeze [16]. Here, we analyzed the data coming from the single-assessor blind follow-up of trial participants at the age of six years [14]. Primary endpoints included current asthma, defined as parent-reported wheeze in the past 12 months or use of asthma medications in the past 12 months, or both. The second primary endpoint was forced expiratory volume in 0.50 seconds, assessed by spirometry. In our analysis, we explored three different scenarios focusing on the parent-reported outcomes only. The scenarios differed based on the number and type of outcomes and were: 1) three outcomes with high incidence: current asthma, wheeze in the past 12 months, and use of asthma medications in the past 12 months; 2) four outcomes: current asthma, wheeze in the past 12 months, use of asthma medications in the past 12 months, and nocturnal cough; 3) four outcomes: current asthma, wheeze in the past 12 months, use of asthma medications in the past 12 months, and use of oral corticosteroids.

### R package *PERMEATE*

We developed an R package, *PERMEATE* (PERmutation basEd ANalysis of mulTiple Endpoints), that returns the power and type I error rate for all presented methods based on user-defined expected effect sizes, incidence, and correlation among the outcomes. Additional documentation can be found [17].

## Results

### Scenario A) MAKI-type data: high and comparable effect size and incidence

When incidence was high and effect sizes were comparable for all the outcomes (similar to the Maki trial), *varP* strongly outperformed both *bonfT* and *minP* for low-to-medium correlation values (*ρ* =0.01 to *ρ*=0.60). When there was high correlation between the outcomes (i.e., *ρ*=0.80), *varP* was still superior, though the difference was reduced compared with the *minP* method (**Figure 2A**). The *minP* approach was superior compared to the *bonfT* across all levels of correlations. The type I error fluctuated around 5% for *varP*, while it was consistently under 5% for *bonfT* and *minP* for all correlation values (**Figure 2B**).

**Figure 1:**
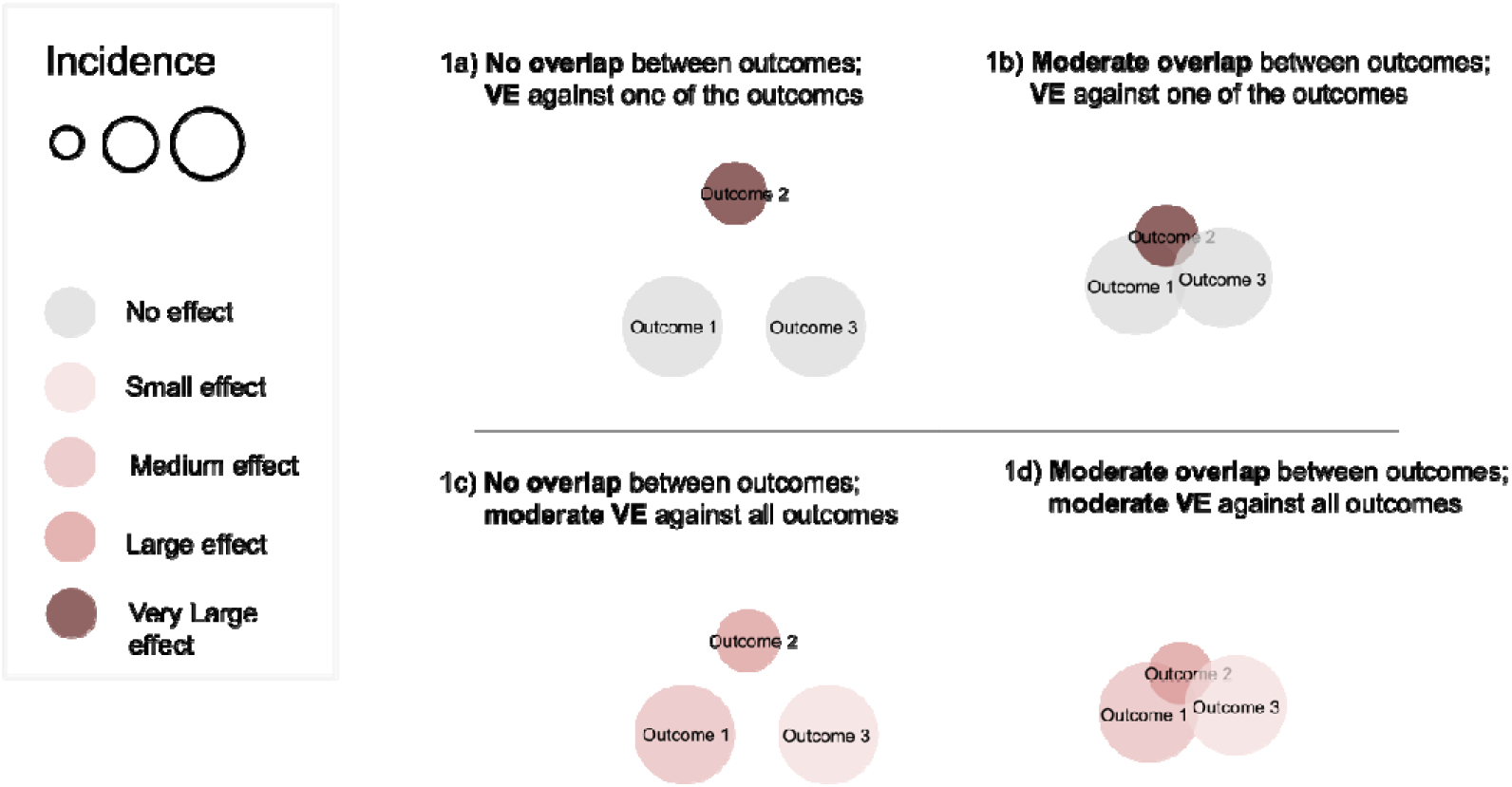
Multiple endpoint scenarios. Four different scenarios are illustrated to represent interactions between clinical outcomes. Panel 1a) shows three independent outcomes with no correlation among them, and there is vaccine efficacy (VE) against one of the outcomes only (outcome 2 in brown); panel 1b) has moderate correlation between the outcomes, and there is VE against one of the outcomes only (outcome 2 in brown); panel 1c) has no correlation between the outcomes, and VE is moderate against all of the outcomes; panel 1d) has moderate correlation between the outcomes, and VE is moderate against all of the outcomes.

**Figure 2:**
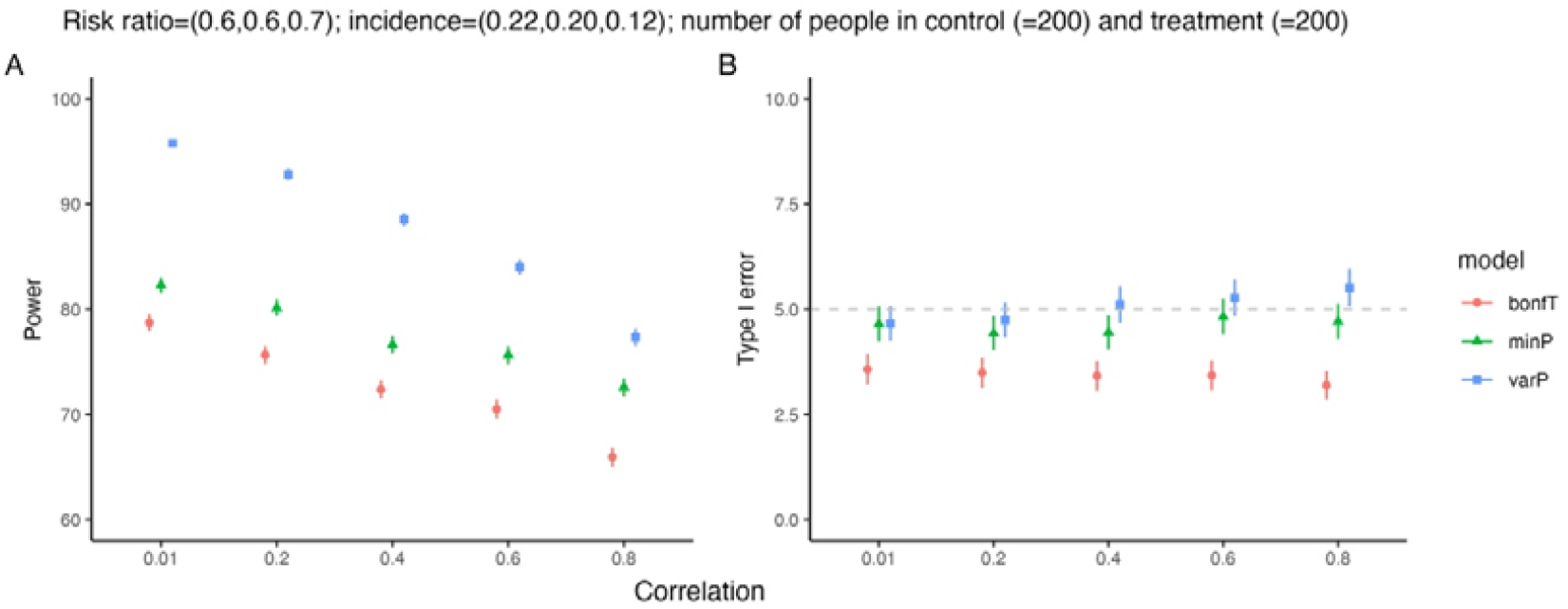
Scenario A) MAKI-type data. Power (panel A) and Type I error (panel B) with varying degrees of correlation among the three outcomes with *X*_*c*_ = *X*_*t*_ = 200, RR=(0.60, 0.60, 0.70), and, *λ*_*c,j*_ =(0.22, 0.20, 0.12). Error bars represent the 95% confidence intervals; *varP* and *minP* are the permutation methods, *bonfT* is the Bonferroni approach; RR= risk ratio;, *λ*_*c,j*_=incidence in the control group for outcome *j*; *X*_*c*_ = number of individuals in the control group; *X*_*t*_ = number of individuals in the treatment group.

### Scenario B) Melody-type data: low incidence and heterogeneous effect size

When the incidence was low and the effect size was heterogeneous among the outcomes (similar to the Melody trial), *varP* exhibited greater power compared to *minP* and *bonfT* in low-correlation scenarios (*ρ*=0.01 to *ρ*=0.40), although the power gains were modest. As the correlation among outcomes increased (*ρ*=0.60), the three approaches had comparable power, while in a high correlation setting (*ρ*=0.80), the power from *varP* was slightly lower than that from *minP* and *bonfT*, but power was still >95% for all methods (**Figure 3A**). The *minP* approach showed modest power gains compared to *bonfT* in all settings. The type I error was around 5% for *varP* and *minP*, but was below the 5% threshold for *bonfT* (**Figure 3B**).

**Figure 3:**
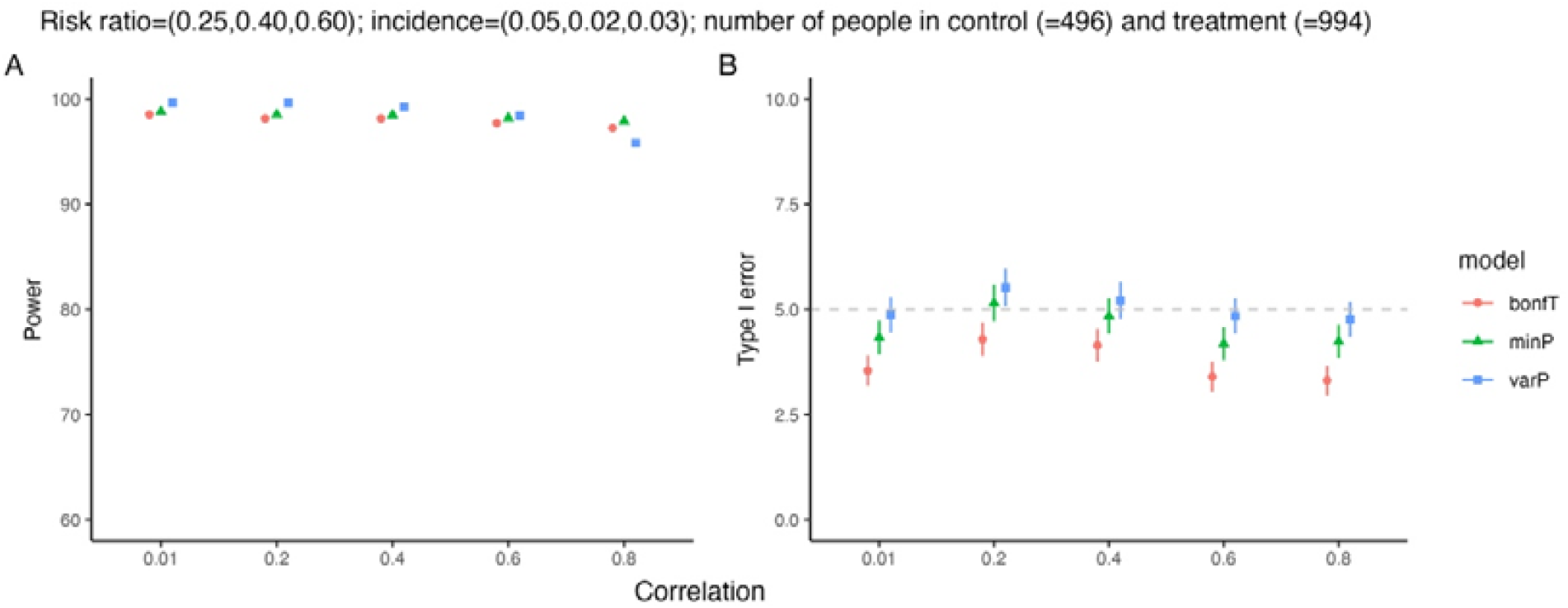
Scenario B) Melody-type data. Power (panel A) and Type I error (panel B) with varying degrees of correlation among the 3 outcomes with *X*_*c*_ = 496, *X*_*t*_ = 994, RR=(0.25, 0.40, 0.60), and, *λ*_*c,j*_ =(0.05, 0.02, 0.03). Error bars represent the 95% confidence intervals; *varP* and *minP* are the permutation methods, *bonfT* is the Bonferroni approach; RR= risk ratio; *λ*_*c,j*_ =incidence in the control group for outcome *j*; *X*_*c*_ = number of individuals in the control group; *X*_*t*_ = number of individuals in the treatment group.

### Scenario C) Prepare-type data: low incidence and high effect size

Finally, in a setting where incidence was low and the rarest outcome had the strongest effect size (similar to the Prepare trial), *varP* outperformed both *minP* and *bonfT* for all correlation values, with a small loss of power as the correlation among the outcomes increased (**Figure 4A)**. The *minP* and *bonfT* approaches showed similar performances, with *minP* slightly superior for all correlation values. The type I error for *varP* fluctuated around 5%, while it was below the threshold for both *minP* and *bonfT* (**Figure 4B**).

**Figure 4:**
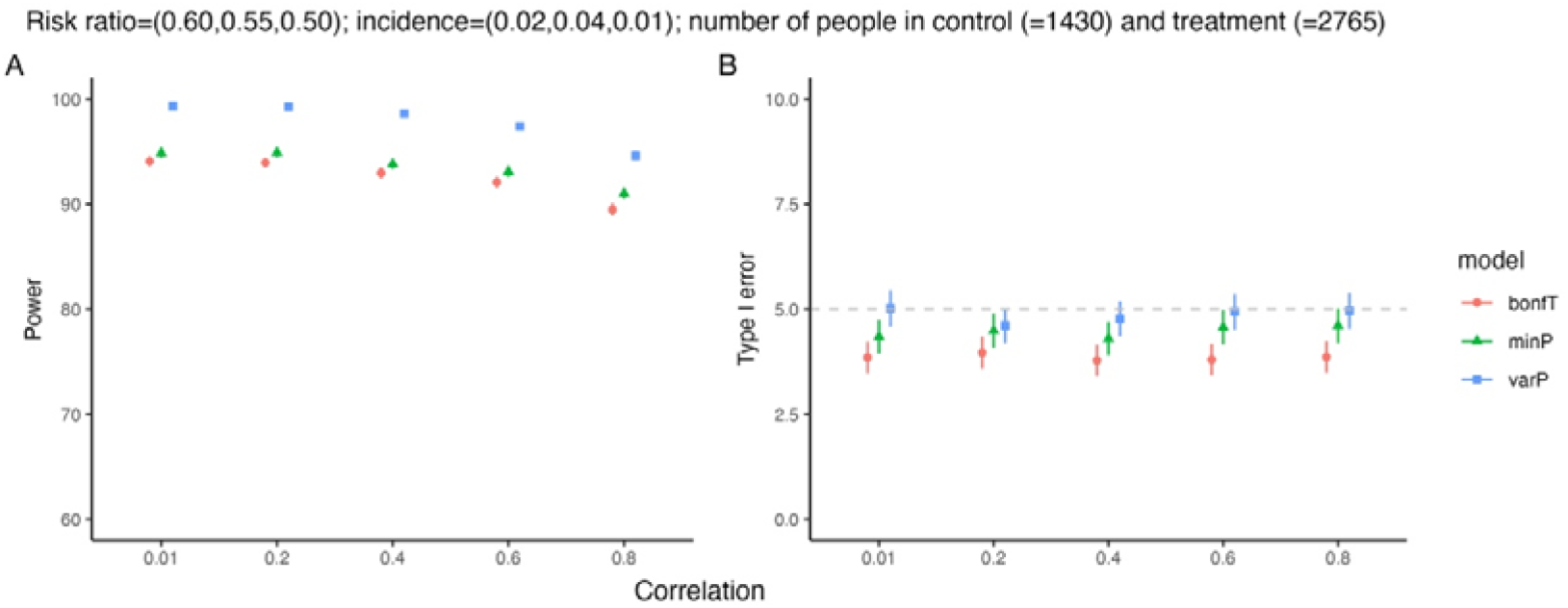
Scenario C) Prepare-type data. Power (panel A) and Type I error (panel B) with varying degrees of correlation among the 3 outcomes with *X*_*c*_ = 1430, *X*_*t*_ = 2765, RR=(0.60, 0.55, 0.50), and *λ*_*c,j*_ =(0.02, 0.04, 0.01). Error bars represent the 95% confidence intervals; *varP* and *minP* are the permutation methods, *bonfT* is the Bonferroni approach; RR= risk ratio; *λ*_*c,j*_ =incidence in the control group for outcome *j*; *X*_*c*_ = number of individuals in the control group; *X*_*t*_ = number of individuals in the treatment group.

For all three simulated scenarios, we also computed the power and type I error when testing each outcome independently, as it would be in a trial with a single endpoint (*Out1, Out2*, and *Out3* for each scenario). In general, the outcome with the highest incidence had the highest power when testing the outcomes independently (**Figures S2 to S4**). When comparing the independent results to a multiple endpoint analysis, *varP* had greater power or similar power to the best performing individual outcome. *varP* outperformed all approaches in scenarios A and C (**Figures S2 and S4**). In scenario B, the power was comparable for *minP, bonfT, varP*, and when looking at *Out2* alone, with a small loss of power for *varP* when correlation was high among the outcomes (**Figure S3**).

### The MAKI trial case study: RSV prevention and asthma in healthy preterm infants

When we analyzed data on three outcomes from the MAKI trial, we found that all the methods returned significant P-values (*p*<0.05). When adding an outcome with high incidence but almost no effect size (i.e., nocturnal cough) to the previous scenario, all methods again had significant P-values. Similarly, when adding a rare outcome with high effect size (i.e., use of oral corticosteroids), all methods had P-values under the 0.05 threshold. In summary, all methods performed similarly in an analysis of this specific real-world dataset. Results are displayed in **Table 2**.

**Table 2:** Analysis of multiple endpoints from the MAKI trial. P-values for the three analytical approaches based on different multiple endpoints evaluated for the MAKI trial data., *λ*_*c,j*_=incidence of outcome *j* in the control arm; *RR*_*j*_= risk ratio for the effect of the intervention (palivizumab for RSV prophylaxis) on each outcome *j, j*=1,..,*M*.

| Description | Selected endpoints<br>(Incidence $\lambda$ and effect size RR) | Analytical approach | P-value |
| --- | --- | --- | --- |
| Three outcomes with comparable incidence and effect size | 1. Current asthma<br>($\lambda_{c,1}=0.24, RR_1=0.59$)<br>2. Wheeze<br>($\lambda_{c,2}=0.20, RR_2=0.58$)<br>3. Use of asthma medications<br>($\lambda_{c,3}=0.13, RR_3=0.71$) | $minP$ | 0.01 |
| | | $varP$ | 0.01 |
| | | $bonfT$ | <0.01 |
| Three outcomes with comparable incidence and effect size, and an additional outcome with high incidence and low effect size | 1. Current asthma<br>($\lambda_{c,1}=0.24, RR_1=0.59$)<br>2. Wheeze<br>($\lambda_{c,2}=0.20, RR_2=0.58$)<br>3. Use of asthma medications<br>($\lambda_{c,3}=0.13, RR_3=0.71$)<br>4. Nocturnal cough<br>($\lambda_{c,4}=0.29, RR_4=0.91$) | $minP$ | 0.02 |
| | | $varP$ | 0.03 |
| | | $bonfT$ | <0.01 |
| Three outcomes with comparable incidence and effect size, and an additional outcome with low incidence and high effect size | 1. Current asthma<br>($\lambda_{c,1}=0.24, RR_1=0.59$)<br>2. Wheeze<br>($\lambda_{c,2}=0.20, RR_2=0.58$)<br>3. Use of asthma medications<br>($\lambda_{c,3}=0.13, RR_3=0.71$)<br>4. Use of oral corticosteroids<br>($\lambda_{c,4}=0.04, RR_4=0.49$) | $minP$ | 0.01 |
| | | $varP$ | <0.01 |
| | | $bonfT$ | <0.01 |

## Discussion

In this study, we evaluated the performance of three different methods for estimating efficacy from vaccine trials that include multiple overlapping or correlated endpoints. We focused on settings where no clear endpoint could be specified *a priori*, and the choice of the endpoint could determine the success or failure of the trial. We tested different methods for addressing multiple endpoints to provide practical recommendations in diverse scenarios based on the characteristics of the outcome and intervention, including incidence and effect size. In line with previous work [2, 11], we found that the use of multiple endpoints can lead to a better performance in terms of power, while keeping the type I error rate controlled, especially when there is no clear primary endpoint. Additionally, we proposed a novel weighted permutation approach, *varP*, that pools the outcome’s test statistics together using a weighted average based on the inverse variances of the estimators. Using simulated and real-world data from clinical trials of RSV interventions, we show that our approach outperforms alternative methods in settings where the effect sizes among the outcomes are related. This result is particularly relevant in RSV clinical trials, where multiple endpoints are often nested within one another, and the overall value of the vaccine can be demonstrated by combining them together. Important examples of correlated outcomes are current wheeze and asthma; or hospitalization from RSV LRTI and RSV LRTI with severe hypoxemia.

Our analysis showed that methods such as *bonfT*, which treat each outcome independently, were robust in settings in which the intervention had a higher effect size against one outcome compared to the others (Figure 3); however, this method consistently exhibited lower power compared to other methods and was overly conservative with regards to type I error. Previous work demonstrated a superiority in power performances of methods that account for the correlation between outcomes, like permutation-based methods such as *minP* [2, 3, 10, 18]. We found that *minP* again performed well in settings where the intervention had a higher effect size against one outcome compared to the others. However, *minP* performed less optimally when the intervention had comparable effect size against the outcomes. For this reason, we developed a novel permutation approach, *varP*, that pools the test statistics of the different outcomes together. This method uses an inverse-variance weighted average of the RRs (on the log scale) to combine the multiple endpoints such that outcomes with low uncertainty receive more weight. Our findings suggest that when the outcomes are related (Figures 2, 4), *varP* outperforms the alternative methods. Finally, we tested the performances of each outcome individually. As expected, the outcome with the highest incidence led to the best power performances, though in most scenarios these results were lower compared to *varP*, and comparable with the results from *minP* and *bonfT*, further confirming the need to incorporate multiple endpoints in the analysis, when the outcomes are related (**Figures S2 to S4**).

Given the large number of RSV vaccines that are in the clinical development pipeline, it is important to have efficient, well-designed studies that can evaluate these alternatives. This will continue to be important as additional vaccines move towards late-stage clinical trials and in trials that might compare the relative effectiveness of different preventive strategies.

Our work has several limitations. First, the simulated scenarios and approaches we explored are not exhaustive of all possible scenarios and of the large literature on multiple endpoint methods [19]. We tested additional alternative approaches, but we found that performances were worse compared to our proposed method. Second, we chose to use *bonfT* as the benchmark because of its simplicity and broad applicability. However, the *bonfT* approach is known to be conservative, especially when there are a large number of positively correlated outcomes, resulting in a decrease of the type I error with a negative impact on the power; more advanced methods to correct for multiple testing have previously been proposed [5].

In our work, we focused on different approaches to deal with multiple and overlapping outcomes. We developed a novel weighted permutation approach, and we compared the performances with other established methods using simulated and real-world data in the RSV field. We demonstrated how this permutation approach should be preferred when outcomes have shared effect size. As a result, we believe our approach is a robust and attractive method for analyzing clinical trial data from RSV products. We provided a case-by-case guidance in our R package *PERMEATE* to help decide the most appropriate method for the design of clinical trials with multiple endpoints.

In conclusion, there is an acute need to design and implement more efficient randomized controlled trials for vaccines. This is needed to maximize the chances that an effective vaccine will receive licensure and will avoid wasting resources in underpowered or inefficient trials. Other novel trial designs could also be considered together with those that evaluate multiple endpoints, including trials that use Bayesian adaptive or sequential designs. The adoption of these novel approaches could speed the introduction of needed products while maintaining rigorous standards of evaluation that are required in clinical trials.

## Data Availability

All data produced are available online at (see link below)

https://github.com/weinbergerlab/Permeate

## Supplementary file

**Figure S1:**
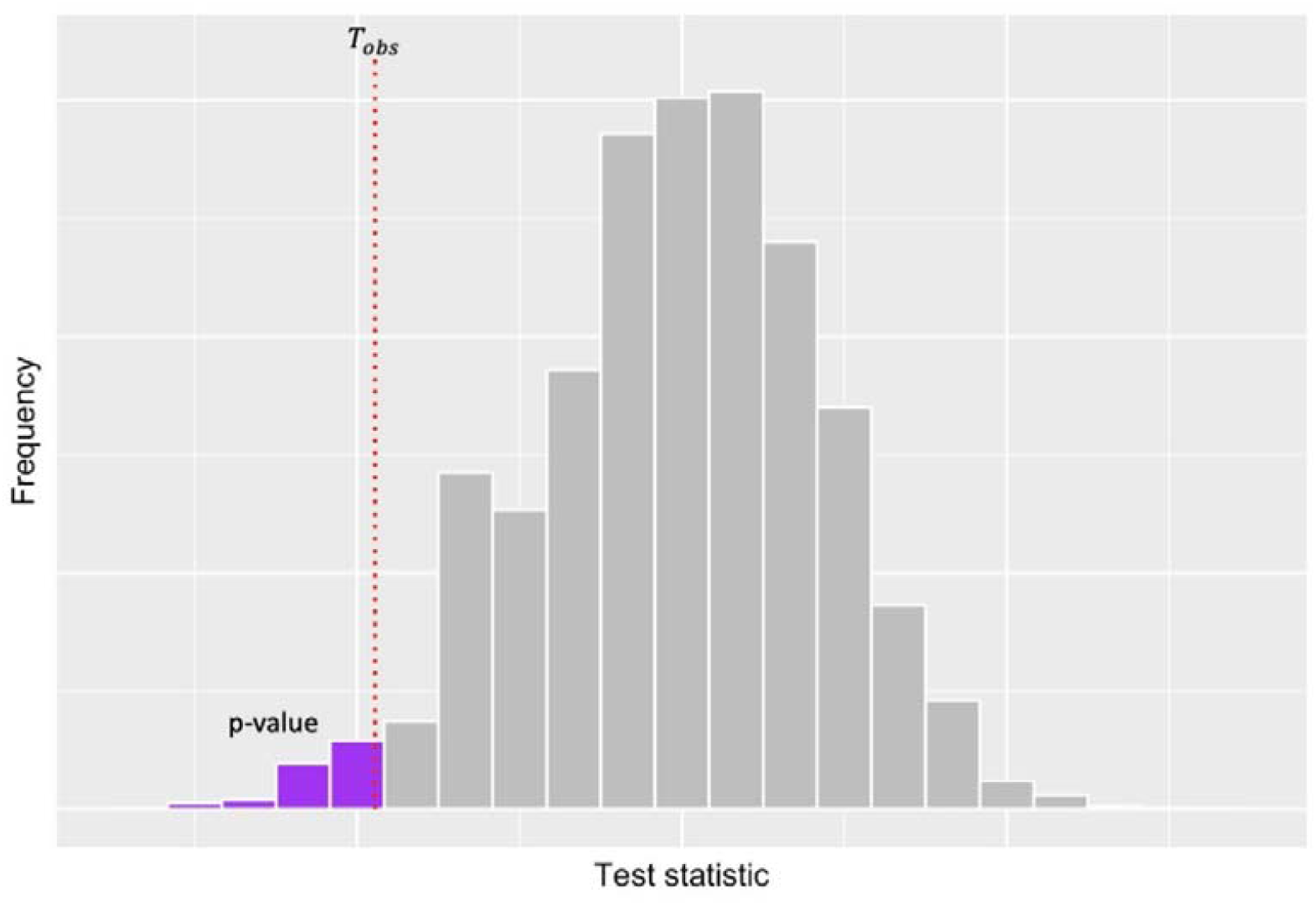
Example of a permutation test. The histogram bars represent the permuted samples (in gray), and the red dotted line corresponds to the observed test statistic. The purple area represents the area where the permuted samples are more extreme than the observed test statistic (i.e., the P-value).

**Figure S2:**
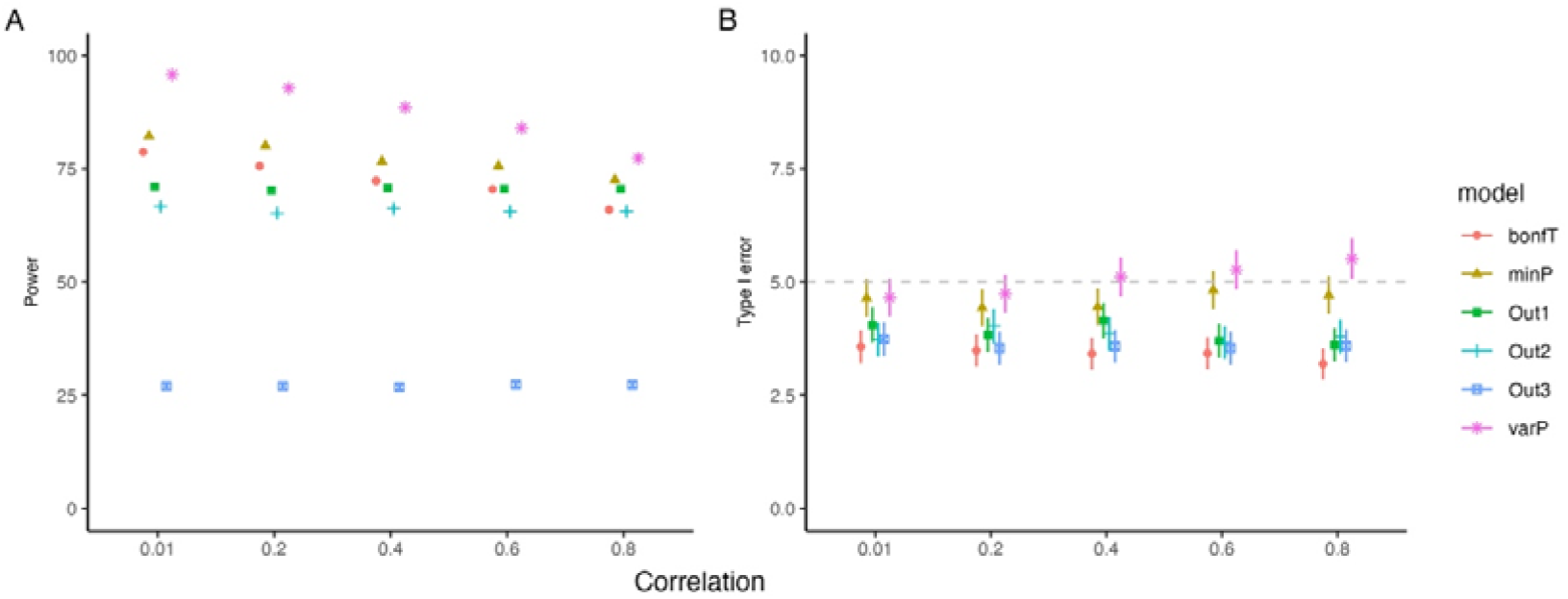
Scenario A) MAKI-type data. Power (panel A) and Type I error (panel B) with varying degrees of correlation among the three outcomes with *X*_*c*_ = *X*_*t*_ = 200, RR=(0.60, 0.60, 0.70), and *λ*_*c,j*_ =(0.22, 0.20, 0.12). Error bars represent the 95% confidence intervals; *varP* and *minP* are the permutation methods, *bonfT* is the Bonferroni approach; *Out1, Out2*, and *Out3* represent the different outcomes analyzed individually; RR= risk ratio; *λ*_*c,j*_ =incidence in the control group for outcome *j*; *X*_*c*_ = number of individuals in the control group; *X*_*t*_ = number of individuals in the treatment group.

**Figure S3:**
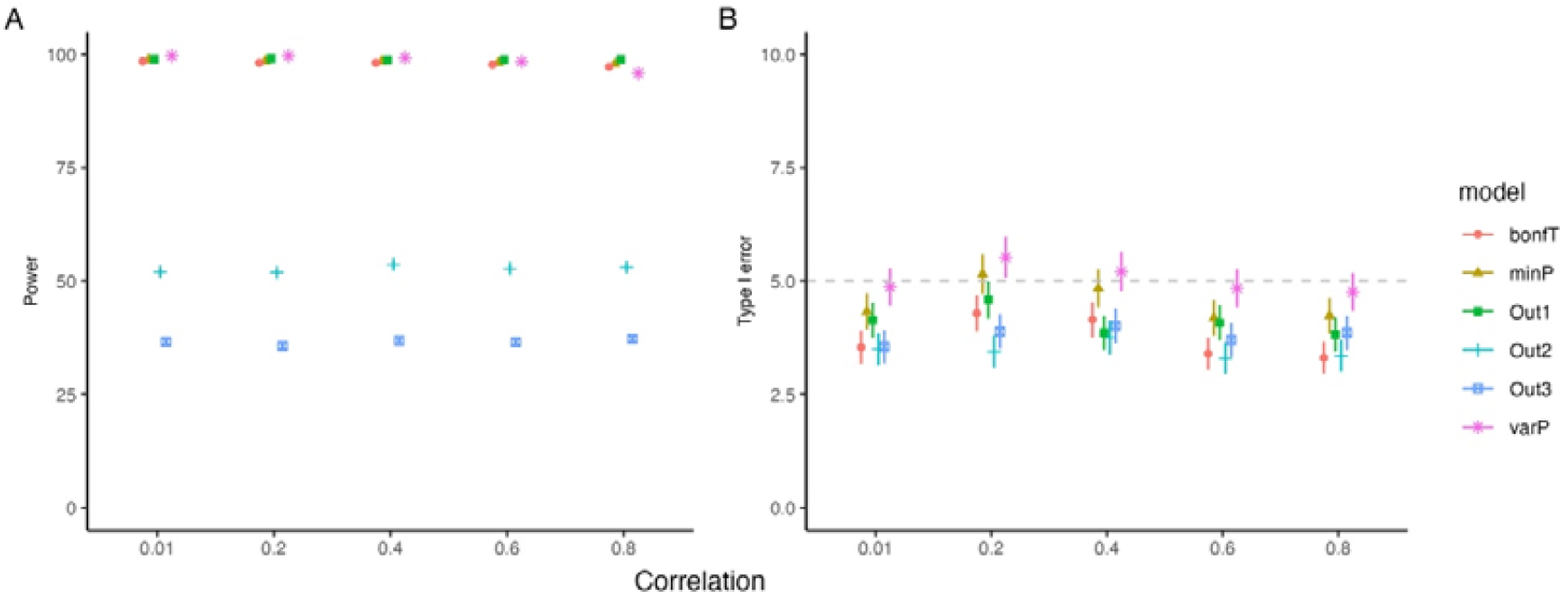
Scenario B) Melody-type data. Power (panel A) and Type I error (panel B) with varying degrees of correlation among the 3 outcomes with *X*_*c*_ = 496, *X*_*t*_ = 994, RR=(0.25, 0.40, 0.60), and *λ*_*c,j*_ =(0.05, 0.02, 0.03). Error bars represent the 95% confidence intervals; *varP* and *minP* are the permutation methods, *bonfT* is the Bonferroni approach; *Out1, Out2*, and *Out3* represent the different outcomes analyzed individually; RR= risk ratio; *λ*_*c,j*_ =incidence in the control group for outcome *j*; *X*_*c*_= number of individuals in the control group; *X*_*t*_ = number of individuals in the treatment group.

**Figure S4:**
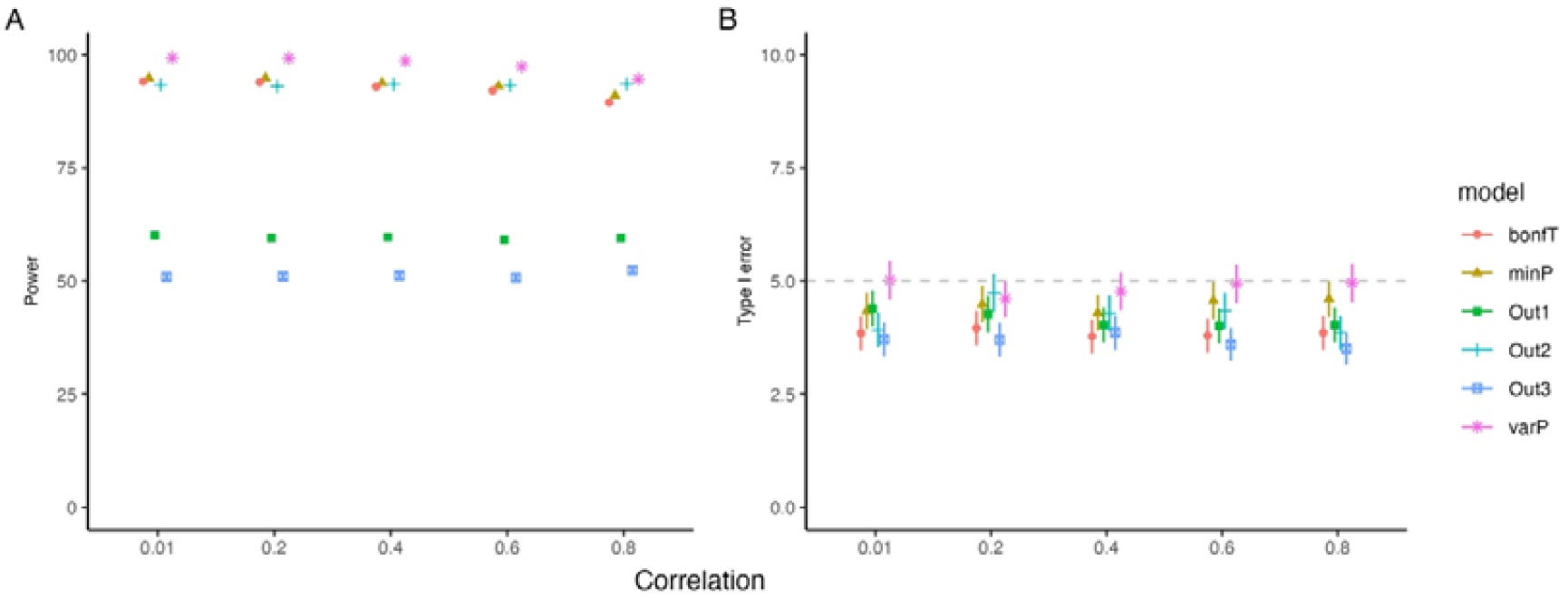
Scenario C) Prepare-type data. Power (panel A) and Type I error (panel B) with varying degrees of correlation among the 3 outcomes with *X*_*c*_ = 1430, *X*_*t*_ = 2765, RR=(0.60, 0.55, 0.50), and *λ*_*c,j*_ =(0.02, 0.04, 0.01). Error bars represent the 95% confidence intervals; *varP* and *minP* are the permutation methods, *bonfT* is the Bonferroni approach; *Out1, Out2*, and *Out3* represent the different outcomes analyzed individually; RR= risk ratio; *λ*_*c,j*_ =incidence in the control group for outcome *j*; *X*_*c*_ = number of individuals in the control group; *X*_*t*_ = number of individuals in the treatment group.

